# Centromedian Nucleus Connectivity with Brainstem Nuclei Unveils a Common Mechanism for Seizure Control

**DOI:** 10.64898/2026.02.18.26346351

**Authors:** Luigi G. Remore, Evangelia C. Tsolaki, Hiroki Nariai, Dawn S. Eliashiv, Aria Fallah, Joyce H. Matsumoto, Noriko Salamon, Marco Locatelli, Ausaf A. Bari

**Affiliations:** Department of Neurosurgery, University of California Los Angeles, Los Angeles, CA, USA; Department of Pathophysiology and Transplantation, University of Milan, Milan, Italy; Department of Neurosurgery, Fondazione IRCCS Ca’ Granda Ospedale Maggiore Policlinico, Milan, Italy; Department of Pediatrics, Division of Pediatric Neurology, University of California Los Angeles, Los Angeles, CA, USA; David Geffen School of Medicine, University of California Los Angeles, Los Angeles, CA, USA; Department of Neurology, University of California Los Angeles, Los Angeles, CA, USA; Department of Radiology, University of California Los Angeles, Los Angeles, CA, USA

## Abstract

**Background:** Epilepsy affects approximately 50 million individuals worldwide, with nearly one-third developing drug-resistant epilepsy (DRE). The centromedian nucleus of the thalamus (CM) and the brainstem are integral components of seizure-modulating networks and represent promising targets for neuromodulation. This study aimed to map structural connectivity between CM and specific brainstem nuclei using probabilistic tractography and to evaluate whether connectivity patterns correlate with seizure reduction following CM-stimulation

**Methods:** Diffusion MRI data from 100 healthy subjects from the Human Brain Connectome database were analyzed to characterize CM–brainstem connectivity. Additionally, 11 patients with generalized DRE who underwent deep brain stimulation (DBS) or responsive neurostimulation (RNS) targeting CM were retrospectively studied. Volumes of tissue activated (VTAs) were used as tractography seeds, and connectivity strength was quantified as probability of connectivity (ProbC), corrected for distance. Two subanalyses were performed by dividing patients into two or three groups based on the threshold for seizure frequency reduction (SFR).

**Results:** In the two-group analysis, responders (>50% SFR) exhibited significantly higher connectivity between VTAs and the nucleus of the solitary tract (NTS) compared with non-responders (<50% SFR), and NTS connectivity was the only parameter significantly correlated with seizure reduction (r=0.762, p<0.001). The three-group analysis confirmed that high responders (>50% SFR) had stronger NTS connectivity than both partial (SFR = 50%) and low responders (<50% SFR), who showed greater connectivity with raphe nuclei.

**Conclusions:** CM–NTS structural connectivity may underlie therapeutic response to CM-neuromodulation, suggesting a potential shared mechanism with vagus nerve stimulation.

## Introduction

Epilepsy is a common neurological condition affecting around 50 million people worldwide, and almost one-third of patients suffer from seizures resistant to pharmacological therapy^1^. The centromedian nucleus of the thalamus (CM) and the brainstem are key components of the neurocircuitry involved in seizure modulation, making them promising targets for neuromodulation therapies aimed at controlling drug-resistant (DRE) epilepsy. CM is a small nucleus, classified in the caudal intralaminar group of thalamic nuclei^2^. In higher primates and humans^3^, CM is histologically distinct from the closely adjacent parafascicular nucleus (Pf) and comprises two subcomponents: a magnocellular part and a parvocellular part, considered the “CM proper”. The first tracing studies in rodents^4^ demonstrated that CM was directly connected to the mesencephalic reticular formation and sent fibers to the basal ganglia, serving as the primary source of extra-cortical afferents to the basal ganglia and participating in sensorimotor regulation. Analogous studies in humans^5^ showed that CM had an enriched pattern of connections with the primary motor and sensory areas, the prefrontal cortex, and the cerebellum. The brainstem is a complex and heterogeneous infratentorial structure, phylogenetically preserved throughout species and classically divided into three anatomical parts: mesencephalon (or midbrain), pons, and medulla oblongata^6^. It is composed of several nuclei and serves as a conduit for asending and descending fibers interconnecting the cortex and many important subcortical structures (cerebellum, thalamus, basal ganglia, and hypothalamus) with the spinal cord and the cranial nerve nuclei with the peripheral efferent ganglia^7^. Along with the most evolutionary archaic functions of autonomic regulation, homeostatic maintenance^8^ and locomotion initiation^9^, the human brainstem is involved in higher brain functions, i.e., arousal and consciousness maintenance^10^, pain regulation^11^ and emotional processing^12^.

Given its widespread connections, neuromodulation targeting the CM using both deep brain stimulation (DBS) and responsive neurostimulation (RNS) has been investigated as an off-label treatment for several neuropsychiatric diseases, such as epilepsy^13^, Tourette syndrome^14^, neuropathic pain^15^, coma, and other vegetative states^16^. For drug-resistant generalized epileptic syndromes, CM-neuromodulation seems to be particularly promising, according to a mean seizure reduction rate of 73.4% after a mean follow-up of 28.7 months as reported by a recent meta-analysis^17^. In this context, we demonstrated in a previous study^18^ that the CM had the strongest structural connections with the brainstem based on a probabilistic tractography analysis conducted on healthy subjects from the Human Brain Connectome (HBC) project. Similar findings resulted from other studies^19,20^, in which different connectomic analyses were performed and a positive correlation between brainstem connections and seizure reduction after CM-DBS was outlined. On the other hand, the role of the brainstem in epileptic syndromes involving loss of consciousness (such as absence epilepsy, focal onset impaired awareness seizures, and generalized tonic-clonic seizures) has been acknowledged since the first electrophysiologic studies in animals^21^ and has been confirmed by modern neuroimaging studies^22^. In fact, decreased activity in the dorsal mesencephalon and upper pons was demonstrated in children with absence seizures during a concomitant EEG-fMRI study^23,24^. However, it is still unclear which brainstem nuclei may play a main role in seizure spreading and if targeted stimulation of the brainstem may help halt epileptic discharges.

In this study, we aimed to map the structural connectivity between CM and the brainstem nuclei using probabilistic tractography. Furthermore, we investigated whether CM connectivity with specific brainstem nuclei is associated with seizure frequency reduction in larger cohort of patients with generalized epilepsy.

## Materials and methods

### Study subjects

We used two distinct subject populations for two different objectives. To map CM structural connections with brainstem nuclei, we analyzed data from 100 unrelated subjects obtained from the Human Brain Connectome (HBC) database^25^ (http://humanconnectome.org). We used the “minimally pre-processed” data with acquisition and pre-processing details previously described ^26^. To investigate the role of CM-brainstem connectivity in seizure frequency reduction, we retrospectively collected clinical and imaging data of 11 generalized DRE patients from our institution’s clinical database. The patients’ cohort comprised two distinct populations: 4 adult patients (≥ 18 years of age) diagnosed with DRE according to the International League Against Epilepsy (ILAE) guidelines^27^, evaluated by neurologists at UCLA Ronald Regan Medical Center and 7 pediatric patients (< 18 years of age) diagnosed by pediatric neurologists at UCLA Mattel Children’s Hospital. Among these patients, 9 underwent CM-RNS and 2 CM-DBS. This study was approved by the Ethical Committee of our institution (IRB #23-000257).

### Patients’ MRI data acquisition and pre-processing

Before surgery, each patient underwent 3T magnetic resonance imaging on a Siemens Prisma scanner, including T1-weighted anatomical images using the MP-RAGE sequence (TE = 2.98 ms, TR = 2.5 s, matrix = 256 × 256, isotropic 1 mm voxels, and flip angle = 15^◦^) and single-shot spin-echo echo planar imaging for diffusion tensor imaging (TE = 91 ms, TR = 7.6s,matrix=128×128,voxelsize=1.7×1.7mm or 2 × 2 mm, slice thickness = 2, b-value = 1000, and 64 directions). MRI scans of pediatric patients were acquired under general anesthesia.

For each patient, T1 images were skull-striped using Brain Extraction Tool (BET)^28^ and non-linearly co-registered to the MNI152 space by using the FNIRT tool in FMRIB Software Library (FSL) (ver. 5.0) (http://fsl.fmrib.ox.ac.uk/fsl)^29^. Eddy current correction was used to apply affine registrations to each volume in the diffusion dataset and register it with the initial reference b0 volume. Before performing tractography, all diffusion data were processed using Bayesian Estimation of Diffusion Parameters Obtained Using Sampling Techniques (BEDPOSTX)^30,31^, to create distributions of diffusion parameters at each voxel. Then, a multi-fiber diffusion model was fitted on HCB data by using the FMRIB’s Diffusion (FDT) toolbox in FSL^31^. This model uses Bayesian techniques to estimate a probability distribution function (PDF) on the principal fiber direction at each voxel, accounting for the possibility of crossing fibers within each voxel. Three fibers were modeled per voxel, a multiplicative factor of 1 for the prior on the additional modeled fibers and 1000 iterations before sampling^30^. Finally, DTI data were linearly co-registered to the structural T1 images with the FLIRT tool and non-linearly co-registered to the standard MNI space as described above.

### Surgical technique

All patients or their legal representatives provided informed written consent for the surgical procedure in accordance with the Declaration of Helsinki. As previously described^18^, our surgical team consisted of two attending neurosurgeons. The CM was targeted indirectly based on stereotactic coordinates as reported in the literature^32,33^ or manually segmented on the T1-weighted image for adult and pediatric patients by an attending neuroradiologist. For all patients, the lead trajectory was planned on the gadolinium-enhanced T1-weighted sequence. Under general anesthesia, a Leksell stereotactic frame (Elekta, Stockholm, Sweden) was placed on the patient’s head, and a thin-layer stereotactic CT was performed. In a dedicated workstation (Elements, BrainLab, Kapellenstrat, Germany), the CT scan was co-registered with MRI sequences containing the stereotactic plan. We bilaterally implanted SenSight directional leads (Model B33005, Medtronic Inc., Minneapolis, Minnesota, USA) for CM-DBS patients and 4-contact depth leads (DL-330-3.5, NeuroPace, Mountain View, California) for CM-RNS patients, respectively. RNS Neurostimulators (model RNS-300M, NeuroPace, Mountain View, California) were placed into a skull mounted tray via a craniotomy during the same surgical procedure, whereas CM-DBS patients had the intracranial electrodes connected to an infraclavicular IPG Percept PC (Medtronic Inc., Minneapolis, Minnesota, USA) in a right hemithorax subcutaneous pouch during a second hospitalization.

### Tractography seed and targets

For HBC subjects, tractography analysis was performed using a CM mask derived from the THOMAS atlas^34^, which was co-registered to each subject’s diffusion space. As target areas, we selected 12 brainstem nuclei taken from three different MNI atlases: the Harvard AAN atlas^10^, the Levinson-Bari atlas^12^, and the DISTAL atlas^35^. Among these 12 nuclei, five were single and located on the midline of the neuraxis, while seven were paired and bilateral (**Figure 1**). All brainstem nuclei masks were non-linearly co-registered to the subject’s diffusion space as described above. For DRE patients, tractography seeds were the volumes of tissue activated (VTAs) computed with the finite element method (FEM)-based FieldTrip SimBio pipeline in Lead-DBS (ver. 2.6)^36^. VTAs were generated based on the stimulation parameters (active contacts and amplitude in mA) recorded at the last follow-up visit. Since no significant side differences were expected in CM connections and the beneficial effect of unilateral CM stimulation has been reported in the literature^33^, right VTAs were mirrored to the left. We then performed probabilistic tractography using the 12 left brainstem targets from the same MNI atlases, increasing the power of our analysis. Each VTA was then co-registered to the patient’s diffusion space before tractography analysis.

**Figure 1.**
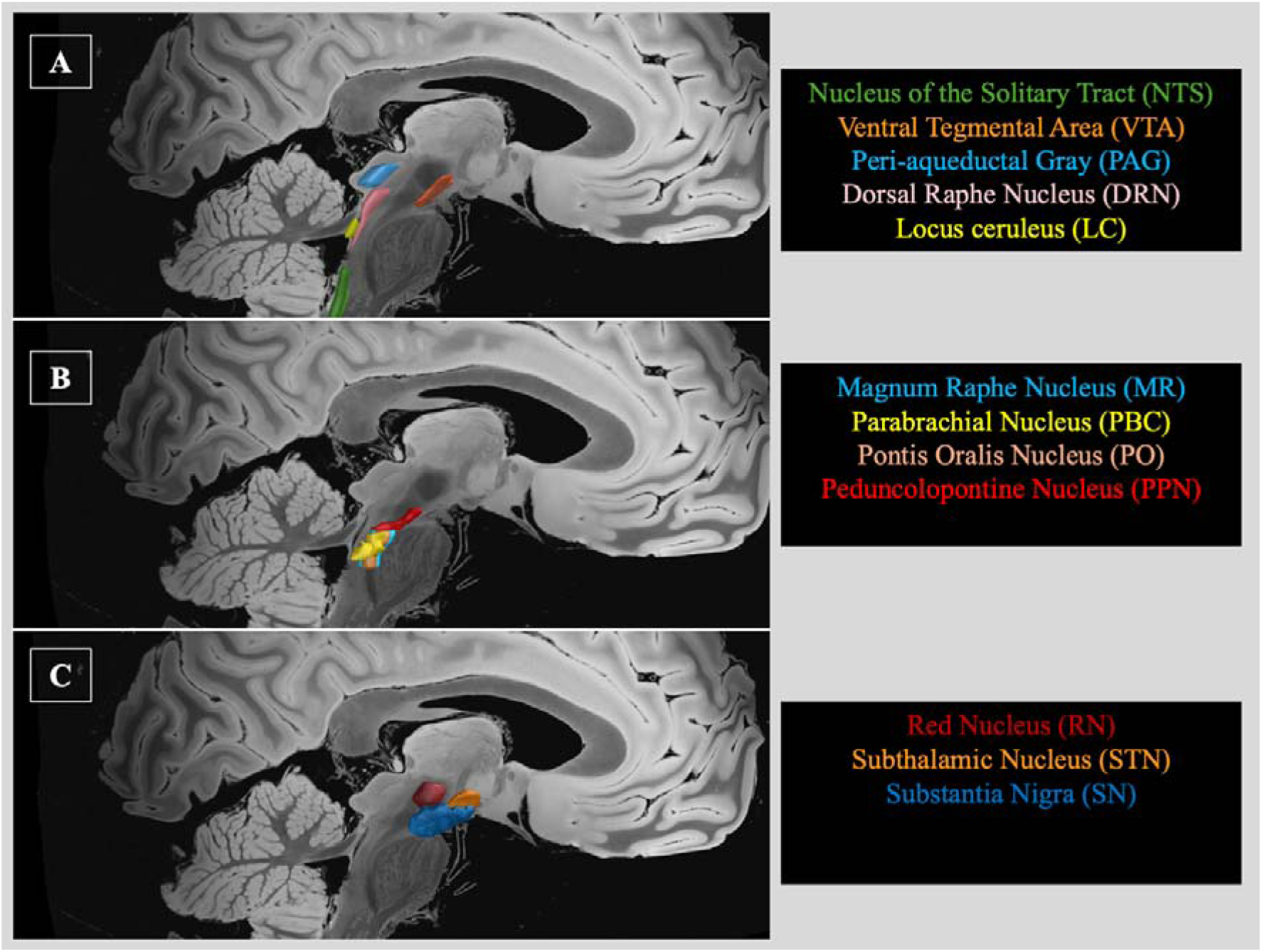
Brainstem nuclei used for the tractography analysis A: Nuclei taken from Levinson-Bari Atlas. B: Nuclei taken from Harvard AAN Atlas. C: Nuclei taken from Distal Atlas.

### Probabilistic tractography

Probabilistic diffusion tractography was performed using the FMRIB’s Diffusion (FDT) toolbox in FSL to define structural connectivity between the brainstem target masks and the seed masks (CM for HBC subjects and VTAs for DRE patients). From each voxel in the three seeds, 5000 streamlines were generated, a 0.2 curvature threshold was chosen, a loop check termination was set and the target masks were used as waypoint masks to discard tracts not passing through the target, termination masks to terminate the pathways as soon as they reach the target and classification masks to quantify connectivity values between the seed and target mask^31^. Then, the probability of connectivity (ProbC) of the seed masks to each target mask was calculated by dividing the number of streamlines connecting the seed to the target by the total number of streamlines sent out (5000 streamlines per seed voxel)^37,38^. To account for the distance-dependent effect, ProbCs values were corrected by the average distance between the seed and the corresponding target masks^39,40^. Therefore, ProbC served as a quantitative metric, representing the strength of structural connectivity between each seed and target.

### Patient’s subgroup analysis

Two subgroup analyses were performed for DRE patients. Firstly, patients were divided into responders and non-responders using a > 50% seizure frequency reduction threshold as a response criterion, and the VTAs’ ProbC values with the brainstem target masks were compared between these two groups. Then, a second analysis was conducted by distinguishing three patient groups: high responders (> 50% seizure frequency reduction), partial responders (exactly 50% seizure frequency reduction), and low responders (< 50% seizure frequency reduction). Neuromodulation for DRE patients is considered a palliative treatment with several approved neuromodulation strategies^41,42^ demonstrating long-term benefits, even below the clinically established 50% response threshold ^43,44^. Therefore, the second subgroup analysis was designed to provide a more detailed evaluation of the actual benefit of neurostimulation on patients’ daily lives. Seizure reduction frequencies were calculated from clinical evaluations conducted by the attending neurologists at the last follow-up visit.

### Statistical analysis

Descriptive statistics were used to summarize demographic characteristics, with frequencies and percentages reported for categorical variables and means ± standard deviations (SD) for continuous variables. A Shapiro-Wilk normality test was used to assess normality across selected variables, along with a close inspection of the data plotted on histograms and Q-Q graphs. Differences in categorical variables were compared with the Chi-Square test.

MANOVA was employed to perform multivariate comparisons among continuous variables between two or more groups. In this context, the Pillai-Bartlett trace (V) multivariate test was used to test for overall statistical significance among all variables, and ANOVA univariate F-statistic was used as a follow-up test in case of multivariate significance. For the patients’ three-group analysis, Bonferroni was used as a post-hoc test to differentiate the directionality of statistical significance among the groups. Finally, Pearson’s correlation (r) was used to examine relationships between two continuous variables. For all traditional hypotheses, p-values < 0.05 were considered statistically significant. Statistical analysis was performed using SPSS (version 29.0, IBM).

## Results

### Connectivity of the centromedian nucleus with the brainstem nuclei

In HBC subjects, CM had the highest ProbC with the parabrachial nucleus (Prob 0.193 ± 0.1) and the weakest ProbC with the subthalamic nucleus (Prob 0.01 ± 0.006) (**Figure 2**) (**Supplementary Table**).

**Figure 2.**
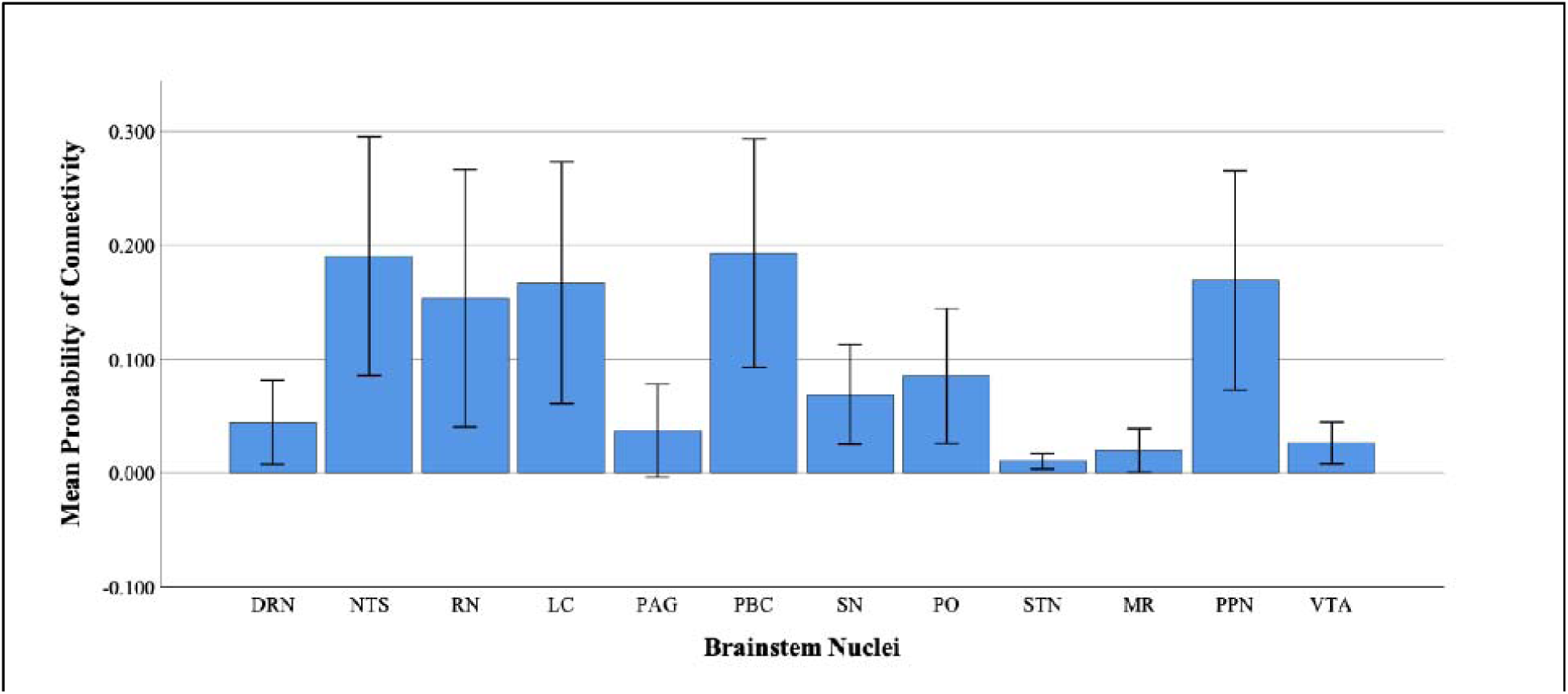
Probability of connectivity of the centromedian nucleus with brainstem nuclei PBC: parabrachial nucleus complex; NTS: nucleus of the solitary tract; PPN: pedunculopontine nucleus; LC: locus ceruleus; RN: red nucleus; SN: substantia nigria; PO: pontis oralis nucleus; DRN: dorsal raphe nucleus; PAG: peri-aqueductal gray; VTA: ventral tegmental area: MR: magnum raphe nucleus; STN: subthalamic nucleus.

### Patient’s clinical characteristics

Eleven patients suffering from generalized DRE were included in the analysis (**Table 1**). 63.3% of the patients (7/11) were males, while the mean age at epilepsy onset was 5.01 ± 5.24 years and the mean age at surgery was 20.02 ± 15 years. Seven patients had previously undergone vagus nerve stimulator (VNS) implantation before CM-neuromodulation, and three patients had received a callosotomy; however, seizures relapsed months after the procedure. Following a mean post-implantation follow-up of 8.55 ± 4 months, the mean seizure frequency reduction rate was 0.6 ± 0.23.

**Table 1:** Clinical characteristics of the patients with chronic generalized epilepsy.

| Sex<sup>1</sup> | Epilepsy/Seizure type | Epilepsy etiology | Stimulation type | Stimulation settings | Two-tier Response<sup>6</sup> | Three-tier Response<sup>7</sup> |
| --- | --- | --- | --- | --- | --- | --- |
| F | Tonic-clonic | Undefined<sup>2</sup> | RNS | Bipolar | Responder | High Responder |
| M | LGS<sup>3</sup> | Genetic | RNS | Bipolar | Responder | High Responder |
| F | Myoclonic | Undefined | RNS | Bipolar | Non-Responder | Low Responder |
| M | LGS | Undefined | RNS | Bipolar | Responder | High Responder |
| M | LGS | Undefined | RNS | Bipolar | Non-Responder | Low Responder |
| M | LGS | Undefined | RNS | Bipolar | Responder | High Responder |
| F | Tonic-clonic | Genetic | RNS | Bipolar | Non-Responder | Partial Responder |
| M | LGS | Cortical dysplasia | RNS | Bipolar | Non-Responder | Partial Responder |
| F | Drop spells | HIE<sup>4</sup> | RNS | Bipolar | Non-Responder | Partial Responder |
| F | JAE<sup>5</sup> | Genetic | DBS | Monopolar | Non-Responder | Partial Responder |
| M | Tonic-clonic | Undefined | DBS | Monopolar | Non-Responder | Low Responder |

1: Patient's sex (M = male, F = female); 2: epilepsy etiology was deemed to be "undefined" after a thorough clinical workout, including careful anamnesis and physical examination, different neuroimaging studies (MRI and PET), video-EEG and stereoEEG in selected cases; 3: LGS = Lennox Gastaut Syndrome; 4: hypoxic-ischemic encephalopathy; 5: JAE = Juvenile Absence Epilepsy. 6: responders = > 50% seizure frequency reduction, non-responders = <50% seizure frequency reduction; 7: high responders = > 50% seizure frequency reduction, partial responders = 50% seizure frequency reduction, low responders = < 50% seizure frequency reduction.

### Two group subanalysis

Four patients were classified as responders, achieving >50% seizure frequency reduction at the last follow-up visit. The multivariate test revealed a statistically significant difference among the clinical variables between responders and non-responders (V=0.82, F(3,7)=10.87, p=0.007). However, univariate follow-up analysis identified the mean reduction in seizure frequency as the only statistically significant difference between the two groups (0.87 ± 0.18 VS 0.45 ± 0.05, p<0.001) (**Table 2**). When comparing the ProbCs values of VTAs with brainstem nuclei, the multivariate test showed a statistically significant difference (V=0.91, F(7,14)=5.53, p=0.015). Nevertheless, the follow-up univariate analysis confirmed that the only significant difference between responders and non-responders was the VTA to the nucleus of the solitary tract (NTS) (0.28 ± 0.13 VS 0.006 ± 0.01, p<0.001) (**Table 3**). Moreover, no differences between the two groups were found in the rate of VNS implantation (p=0.48) and callosotomy (p=0.9) (**Figure 3A**).

**Figure 3.**
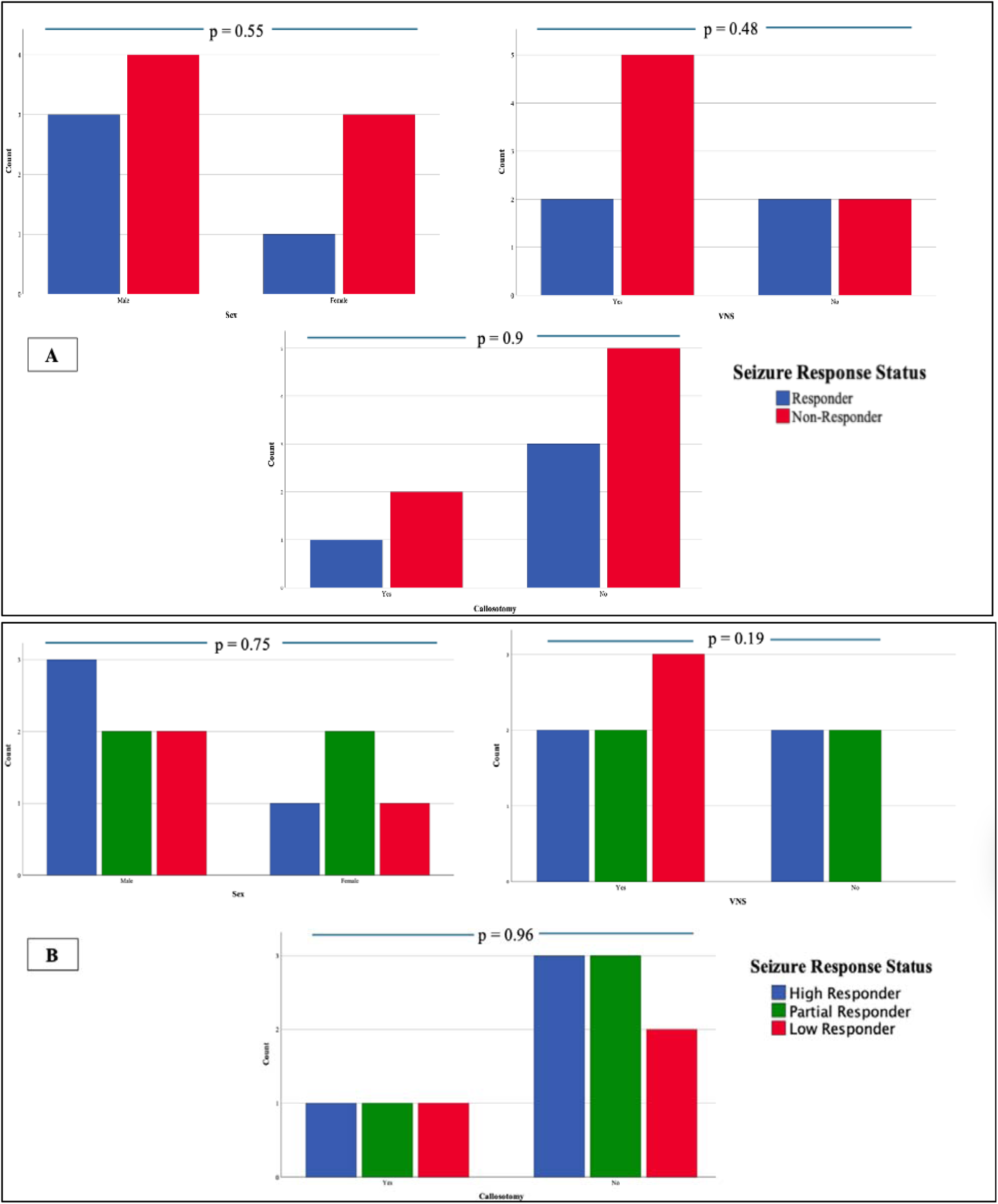
Clinical characteristics of patients in the two subanalyses. Absolute count of patients for the three dichotomic variables: sex, callosotomy, and VNS (vagus nerve stimulation) implantation. P-values refer to the results of the Chi-Square test. A: Two-group subanalysis. B: Three-group subanalysis.

**Table 2.** Patient’s clinical characteristics for the two subgroups analysis.

| Variable | Groups<sup>1</sup> |  |  | F<sup>2</sup> | p-value<sup>3</sup> |
| --- | --- | --- | --- | --- | --- |
|  | Responders | Non-responders |  |  |  |
| Age at onset<sup>4</sup> | 2.81 ± 3.5 | 6.27 ± 5.88 |  | 1.12 | 0.31 |
| Age at implantation<sup>5</sup> | 22.25 ± 25.72 | 18.75 ± 6.02 |  | 0.12 | 0.73 |
| Seizure reduction<sup>6</sup> | 0.87 ± 0.18 | 0.45 ± 0.05 |  | 32.55 | <0.001 |
|  | High Responders | Partial responders | Low responders |  |  |
| Age at onset | 2.81 ± 3.5 | 7.41 ± 6.2 | 4.75 ± 6.3 | 0.73 | 0.5 |
| Age at implantation | 22.25 ± 25.72 | 21.2 ± 6.61 | 15.5 ± 4 | 0.16 | 0.85 |
| Seizure reduction | 0.87 ± 0.18 | 0.5 | 0.4 | 17.55 | 0.001 |

1: Patient's subgroups (see the text for further detail); 2: F-statistics and p-value reported from comparison among probabilities of connectivity by using ANOVA; 3: p<0.05 was considered statistically significant; 4: patient's age at seizures onset expressed in years; 5: patient's age at implantation expressed in years; 6: percentage of seizure frequency reduction expressed in continuous number as recorded by the neurologist during the last follow-up visit.

**Table 3.**
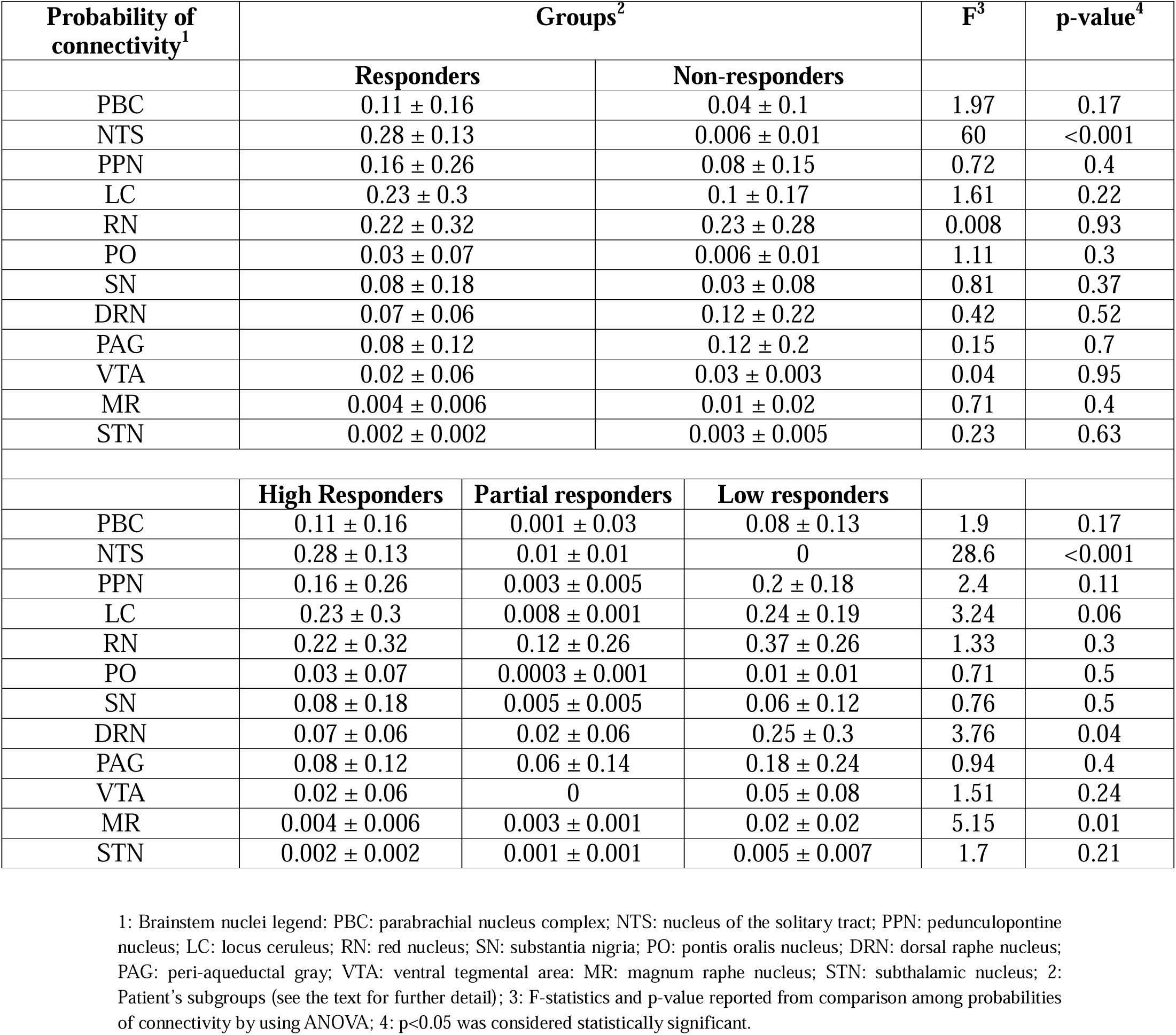
Comparison of the probability of connectivity with target brainstem nuclei in the two subanalyses.

### Three group subanalysis

The distribution of patients was as follows: four high responders, four partial responders, and three low responders. Comparing clinical variables among the three groups, the multivariate test was statistically significant (V=0.9, F(3,7)=12.65, p=0.003), but the univariate test again confirmed that the only statistically significant difference resided in the mean reduction in seizure frequency (p=0.001) (**Table 2**). Specifically, the post-hoc test clarified that high responders had greater mean seizure reduction frequency than both partial responders (p=0.005) and low responders (p=0.002), while no differences (p=0.84) resulted between partial and low responders. Similar to the previous analysis, the rates of VNS implantation (p=0.19) and callosotomy (p=0.96) were not statistically different among the three patient groups (**Figure 3B**).

From the comparison of VTAs’ ProbCs with brainstem nuclei, the multivariate test was statistically significant (V=1.68, F(14,28)=2.61, p=0.031), and the univariate test showed significant differences in the ProbCs with the following nuclei: NTS (p<0.001), dorsal raphe nucleus (DRN) (p=0.042) and magnum raphe nucleus (MR) (p=0.016) (**Table 3**). Post-hoc analysis revealed that high responders had higher ProbC with NTS (0.28 ± 0.13) than both partial responders (0.01 ± 0.005, p<0.001) and low responders (0, p<0.001), while no significant differences were observed between partial and low responders (p=0.9). For DRN connectivity, low responders had significantly higher ProbC than partial responders (0.25 ± 0.3 VS 0.02 ± 0.06, p=0.04). However, no significant differences were found between low responders and high responders (0.07 ± 0.06, p=0.142) and between high and partial responders (p=0.9). Regarding MR connectivity, low responders presented stronger structural connectivity to MR (0.02 ± 0.03) than both high responders (0.004 ± 0.006, p=0.05) and partial responders (0.003 ± 0.001, p=0.02), while no significant differences were observed between high and partial responders (p=0.9). Nonetheless, the ProbC with NTS was the only connection directly correlated with changes in seizure frequency (r = 0.762, 95% CI [0.5, 0.9], p<0.001) (**Figure 4**).

**Figure 4.**
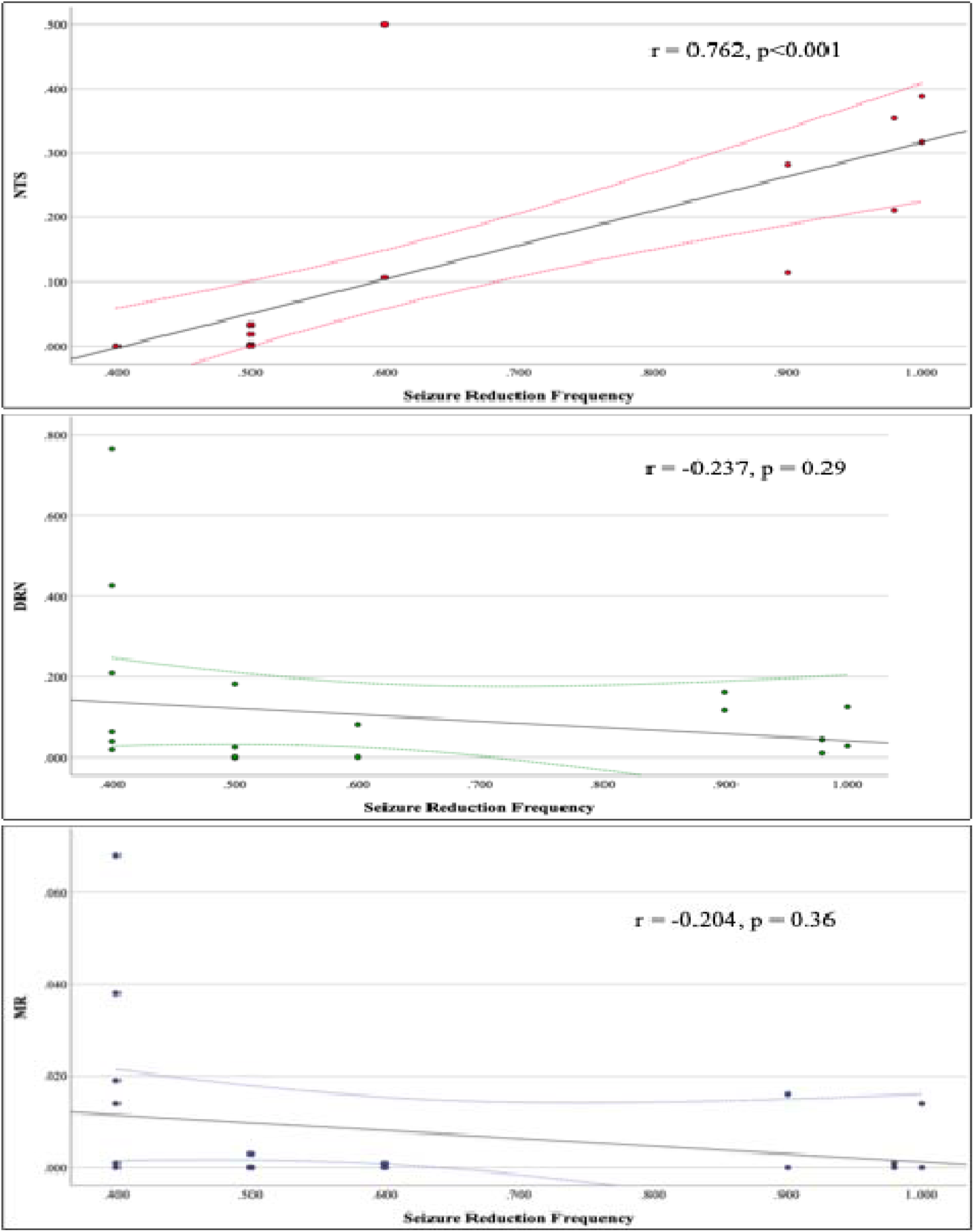
Correlations between seizure reduction frequency and probability of connectivity with brainstem nuclei in patients. NTS: nucleus of the solitary tract. DRN: dorsal raphe nucleus. MR: magnum raphe nucleus.

## Discussion

In this probabilistic tractography study, we show that the CM nucleus has important structural connections with several brainstem nuclei, particularly those in the mesencephalon and upper pons. This finding is consistent with our previous work^18^, in which we used the same CM mask from the THOMAS atlas as seed and a whole brainstem mask as the target. The rostral brainstem has long been recognized as a critical region for arousal and consciousness regulation, dating back to the classic experiment by Moruzzi and Mogun^45^, in which stimulation of the dorsal midbrain and pontine reticular formation in anesthetized cats produced desynchronized EEG activity. Conversely, lesions in the same anatomical area resulted in coma with synchronized EEG. The anatomo-physiological basis of this phenomenon was correlated with the direct connections of the brainstem reticular formation with the intralaminar reticular nuclei and the consequent widespread activation of the neocortex^46^. Given that the CM nucleus is located in the intralaminar thalamic area^2^ and that loss of consciousness is a common feature in many generalized epileptic syndromes^22,23^, our findings support the hypothesis that the anti-seizure effect of CM-neuromodulation may also be mediated by direct connectivity with the brainstem.

Among the brainstem nuclei, CM exhibited the highest probability of connectivity with the parabrachial nucleus (PBC), a three-component complex consisting of lateral, medial, and subparabrachial nuclei^47^. The PBC is particularly significant due to both its anatomical location and structural connections. It plays a crucial role in autonomic regulation, including respiration^48^, blood pressure, water homeostasis^49^, and thermoregulation^50^, as well as in sensory processing, such as gustatory^51^ and modulation of painful stimuli ^52^. More recently, it has also been shown to be involved in emotional processes^53^. PBC receives nociceptive and visceral afferents from the NTS, but it is also directly connected with the Purkinje neurons of the cerebellum^54^. Additionally, it sends efferent projections to several brain areas, including the intralaminar thalamic nuclei, the hypothalamus, the basal forebrain^55^ and multiple cortical and subcortical limbic hubs, such as the amygdala, hippocampus, bed nucleus of the stria terminalis, anterior insular cortex, anterior cingulate cortex, and the orbitofrontal cortex^56–59^. This latter connectivity is especially notable considering our previous study^18^, which unexpectedly found that CM’s direct structural connections with limbic structures were the weakest among all the analyzed target areas. While some studies speculate that limbic structures were more related to seizure spreading and maintenance than generation in generalized epileptic syndromes^60^, the role of the limbic system in epilepsy remains critical. The current findings suggest that CM’s connectivity to PBC may serve as an indirect route to multiple limbic structures, potentially contributing to its therapeutic effect in epilepsy as well as in other pathologies with a prominent limbic substrate, i.e., Tourette syndrome^14^ and neuropathic pain^15^.

Subgroup analysis revealed that the ProbC between VTAs and NTS was significantly higher in responders than in non-responders and was the only connection statistically correlated with seizure-frequency reduction. Moreover, patients displaying less than 50% seizure reduction exhibited no VTA connections at all with the NTS in the three-group subanalysis. The NTS is a large nucleus extending along the craniocaudal axis in the medulla, receiving nociceptive and visceral afferents from the facial, glossopharyngeal, and vagus cranial nerves. It sends efferents above all to the PBC, although NTS projections to the LC and DRN are also demonstrated^12^. Along with the well-known role in the autonomic response to visceral stimuli from the cardio-pulmonary and gastrointestinal systems^61^, the NTS has direct connections with the amygdala and the dorsolateral prefrontal cortex and has been implicated in fear processing and memory consolidation^62^. Interestingly, the NTS is at the base of the putative mechanism of action of the cervical vagus nerve stimulation (VNS), which is an alternative neuromodulation treatment for drug-resistant epilepsy^63^. Since NTS receives over 75% of all vagal afferent fibers, VNS is thought to exert its anti-epileptic effects via NTS direct connections with the PBC, LC, and DRN, which, in turn, relay to the thalamus, the amygdala, the hippocampus, and several cortical areas^64^. Although it is traditionally believed that the NTS had indirect connectivity with the thalamus via the other brainstem nuclei, more recent evidence suggests the existence of direct NTS-thalamus connections. In rats, retrograde viral track tracing studies show direct connectivity between the NTS and Pf, a pathway linked to gastrointestinal sensations evoked during temporal lobe seizures^65^. In primates, tracer studies have shown that the rostral (prevagal) segment of NTS projects primarily to the ventro-caudal nucleus of the thalamus, while the caudal (postvagal) segment is connected to the PBC and nucleus ambiguous^66^. Since the CM is not fully distinguishable from Pf in rats and the CM remains in proximity to the sensory thalamus (i.e., the ventro-caudal nucleus complex) in primates, it is feasible that direct CM-NTS connections have developed in humans - though histological confirmation is still lacking. Based on this connectivity, CM may be a more effective target than the vagus nerve, as it directly connects to the NTS along with multiple other brain areas associated with seizure control, such as the motor cortex, the prefrontal cortex, and the cerebellum^67,68^. However, an alternative and potentially more intriguing hypothesis is that CM stimulation may enhance the anti-seizure effect of VNS. In fact, many patients receive VNS before being considered for CM-stimulation, as was the case in our cohort (6/9). Moreover, VNS is typically performed unilaterally on the left side due to anatomical factors (more prominent cardiac fibers in the right vagus) and excessive bradycardia observed during right-sided VNS in pre-clinical studies in dogs^69^. Thus, concurrent CM and VNS stimulation could potentially activate the NTS-PBC network bilaterally, leading to a broader anti-epileptic effect. Preliminary findings from the ADVANCE (Add-on Deep brain stimulation versus continued VAgus Nerve stimulation for Childhood Epilepsy) study support this hypothesis. In this prospective, non-blinded, randomized patient preference trial, 18 children displayed higher seizure control with DBS as add-on therapy after at least one year of insufficient seizure control with VNS only^70^.

On the other hand, low responder patients had a higher probability of connectivity with the DRN and MR, both of which are components of the raphe nuclear complex. This complex is classically divided into two groups^71^: the rostral group (midbrain raphe), which includes three nuclei (DRN, median raphe nucleus, caudal linear nucleus) and is involved in sensory and emotional processing and regulation of food intake and the circadian rhythm^72^; the caudal group (medullary raphe), which includes three nuclei (MR, raphe pallidus nucleus and raphe obscurus nucleus) and plays a role in autonomic and somatosensory responsiveness, especially for respiration and pain control^73,74^. The raphe nuclear complex is the major source of endogenous intracerebral serotonin, which has an acknowledged role in epilepsy, especially regarding depression and other mood comorbidities in epileptic patients^75^. Furthermore, serotonin and the medullary raphe have been involved in the pathophysiology of sudden unexpected death in epilepsy (SUDEP)^76^. SUDEP accounts for up to 17% of all epilepsy-related deaths and up to 50% of DRE patients^77^. Neurons of the medullary raphe act as central chemoceptors, thereby sensing blood increases in carbon dioxide – a likely occurrence during a prolonged generalized seizure- and activating the respiratory drive in the pons^78^. Notably, several studies have shown that patients who later died of SUDEP had a decreased brainstem volume on structural MRI^79^ and a reduced serotonin receptor binding in the raphe nuclei on PET imaging^80^. Interestingly, the only significant difference between low-responders and the other two patient subgroups was their connectivity with the MR, a key structure of the medullary raphe. This suggests that low responders may have failed to benefit from stimulation due to predominant VTA connectivity with a brain area that is anatomically damaged and strongly associated with SUDEP in DRE patients.

### Limitations

A limitation of this study lies in the intrinsic technical difficulty when analyzing MRI images of the brainstem. Arterial pulsation from major vessels such as the basilar artery may generate motion artifacts that undermine the quality of diffusion images and complicate the identification of the brainstem nuclei, whose borders are anatomically blurred per se^81^. To overcome this issue, we utilized brainstem nuclei masks from externally validated MNI atlases and based our tractography analysis on modern probabilistic algorithms to enhance accuracy. Finally, our patients’ cohort was relatively small, which may have limited the statistical significance of our results to the NTS nucleus only, whereas a larger dataset could have potentially revealed more significant differences across additional brainstem regions.

## Conclusions

Our study demonstrates that the CM has strong structural connections with PBC and NTS. Furthermore, responders exhibited higher VTA connectivity with the NTS, which was the only factor significantly correlated with seizure-frequency reduction. These findings, combined with the established role of the PBC and NTS as key components in the neurophysiological mechanism of VNS, suggest a potential shared anti-seizure mechanism between the two neuromodulation techniques. However, no significant differences were found in VNS implantation rates among the patient groups, indicating that higher NTS connectivity may be necessary but not sufficient for CM-neuromodulation success in DRE patients. Other mechanisms or patient-specific factors not accounted for in this study may contribute to CM stimulation response variability. Therefore, further studies in larger patient cohorts are warranted to investigate the potential additive therapeutic effects of combined CM and VNS stimulation.

## Data Availability

All data produced in the present study are available upon reasonable request to the authors

