## Supplementary Table for "Centromedian Nucleus Connectivity with Brainstem Nuclei Unveils a Common Mechanism for Seizure Control"

**Supplementary Table:** Mean probability of connectivity of the centromedian nucleus with target brainstem nuclei in HBC subjects.

| **Brainstem Nuclei^1^** | **Probability of connectivity** |
| --- | --- |
| PBC | 0.193 ± 0.1 |
| NTS | 0.19 ± 0.1 |
| PPN | 0.17 ± 0.1 |
| LC | 0.16 ± 0.1 |
| RN | 0.15 ± 0.11 |
| PO | 0.08 ± 0.06 |
| SN | 0.07 ± 0.04 |
| DRN | 0.04 ± 0.03 |
| PAG | 0.03 ± 0.04 |
| VTA | 0.02 ± 0.02 |
| MR | 0.01 ± 0.01 |
| STN | 0.01 ± 0.006 |

1: PBC: parabrachial nucleus complex; NTS: nucleus of the solitary tract; PPN: pedunculopontine nucleus; LC: locus ceruleus; RN: red nucleus; SN: substantia nigria; PO: pontis oralis nucleus; DRN: dorsal raphe nucleus; PAG: peri-aqueductal gray; VTA: ventral tegmental area: MR: magnum raphe nucleus; STN: subthalamic nucleus.
